# Mass Spectrometry-Based Amyloid Typing in FFPE Biopsies: A Five-Year Prospective Experience from a Tertiary Academic Center in Spain

**DOI:** 10.64898/2026.10.20.26364269

**Authors:** Valentina Calamia, Patricia Fernández-Puente, Selva Riva-Mendoza, Patricia Quaranta, Beatriz Rocha, Lucía Lourido, Jorge Pombo-Otero, María Generosa Crespo-Leiro, Olalla Ramil-Gómez, Gonzalo Barge-Caballero, Juan Mosquera, Teresa Hermida, Angel Concha, Cristina Ruiz-Romero, Francisco J Blanco

## Abstract

**Background:** Accurate amyloid subtyping is essential for patient management but remains challenging with antibody-based methods. We prospectively evaluated a standardized LC-MS/MS workflow for routine amyloid typing, focusing on diagnostic yield, operational performance, and added value in cases not fully resolved by conventional methods.

**Methods:** From 2020 to 2025, 115 FFPE biopsies from patients with suspected amyloidosis were analyzed at Hospital Universitario A Coruña, Spain, without laser microdissection. Proteins were identified and quantified using PEAKS, with a peak-area cutoff ≥10^5^ for candidate fibril proteins.

**Results:** Seventy-nine cases (69%) were amyloid-positive, 22 (19%) negative and 14 (12%) indeterminate. AL (51%), ATTR (32%) and AA (10%) predominated, with additional AH (5%), mixed ATTR+AL, hereditary ATTRv and AIns amyloidoses. Binary concordance with conventional techniques ranged from 85.7% to 91.5%. LC-MS/MS provided additional diagnostic information in 64/110 evaluable cases (58.2%), including additional subtyping information in 53/110 (48.2%). The workflow also enabled relative quantification of variant versus wild-type TTR peptides and characterization of SAA post-translational modifications. Mean turnaround time was approximately two weeks and MS findings informed clinical management.

**Conclusions:** LC-MS/MS amyloid typing was clinically feasible in routine FFPE biopsies without laser microdissection and supported identification of common, rare and mixed amyloid types.

## Introduction

Amyloidosis comprises disorders caused by extracellular deposition of misfolded protein fibrils. More than 40 amyloidogenic proteins are recognized, including immunoglobulin light chains (AL), transthyretin (ATTR), serum amyloid A (AA), and hereditary or iatrogenic proteins [1,2]. Accurate subtype identification is essential because treatment differs markedly, for example chemotherapy for AL versus TTR-targeted therapy for ATTR [3,4].

Amyloid deposition is conventionally confirmed by Congo red staining and subtyped using immunohistochemistry (IHC) or immunofluorescence (IF) [5,6]. Antibody-based methods may yield inconclusive or misleading results because of cross-reactivity, nonspecific staining, limited validated reagents for rare types, and subjective interpretation [3,7]. Mixed deposits can be particularly difficult to resolve.

Mass spectrometry (MS)-based proteomics has emerged as a reference method for amyloid typing [1,4,8,9]. Typical workflows enrich Congo red-positive deposits by laser microdissection (LMD), followed by protein extraction, digestion, and LC-MS/MS [10]. Sequence-based identification enables simultaneous detection of candidate fibril proteins and universal amyloid-associated proteins such as apolipoprotein E (APOE), apolipoprotein A-IV (APOA4) and serum amyloid P component (SAMP), including rare and hereditary types [1,3,4].

Here, we describe the prospective five-year implementation of LC-MS/MS amyloid typing at Hospital Universitario A Coruña (HUAC), Spain, using routine FFPE sections without LMD. We evaluated 115 consecutive biopsies, diagnostic yield, agreement with conventional methods, turnaround time, and added value in unresolved cases, together with exploratory characterization of sequence variants and post-translational modifications. To our knowledge, this is the first prospective series describing routine LC-MS/MS amyloid typing of FFPE biopsies within an integrated tertiary-hospital pathology/proteomics workflow and the first such experience reported from Spain.

## Materials and Methods

### Patients and samples

A total of 115 tissue biopsies from patients with suspected amyloidosis were analyzed. Specimens were referred from multiple hospital departments including Cardiology, Neurology, Hematology, Nephrology, Pulmonology and Rheumatology and comprised endomyocardial, renal cortical, peripheral nerve, gastrointestinal tract and soft tissue biopsies, reflecting the heterogeneous organ involvement of amyloidosis. All specimens were formalin-fixed, paraffin-embedded (FFPE) tissues. For each case, the Pathology Department provided up to five unstained 10 μm-thick FFPE sections containing the Congo red-positive area. Each analytical batch included an amyloid-negative control tissue of the same histological type (e.g., normal myocardium or renal tissue) for comparative analysis. During the initial phase of the study, positive controls (tissues with histologically confirmed ATTR or AL amyloid deposits) were used to establish the diagnostic thresholds above which a sample was classified as positive.

### Ethics approval, informed consent and data governance

The study protocol, developed in accordance with the Declaration of Helsinki and approved by the Galician Research Ethics Committee. Cardiac biopsies were obtained from patients enrolled in the Galician Cardiac Amyloidosis Registry (AMI-GAL) [11,12]. Clinical source data were accessed through IANUS, the electronic health record system of the Galician Public Health Service, by authorized healthcare personnel. Clinically relevant LC-MS/MS results, once reviewed and validated by the Pathology Department, were incorporated into patients’ electronic health records as part of routine diagnostic care. For research purposes a pseudonymized clinical and analytical dataset was stored on secure institutional systems, with access restricted to authorized members of the study team and processed in accordance with applicable institutional and data-protection regulations.

### Sample processing and protein extraction

Tissue sections were deparaffinized in xylene (2 x 10 min), rehydrated through graded ethanol (100%, 75%), and rinsed in water, with centrifugation between washing steps. Pellets were dried in a SpeedVac concentrator. Proteins were extracted with a strong lysis buffer optimized for amyloid extraction, composed of 20 mM Tris-HCl pH 8.8, 2% SDS and 200 mM DTT. Samples were incubated at 100°C for 20 min with vortexing and then at 80°C for 2 h to reverse formaldehyde crosslinks and solubilize amyloid fibrils. After cooling, lysates were centrifuged (14,000 rpm, 15 min, 4°C) and supernatants were collected. Proteins were precipitated with pre-chilled acetone (6:1 ratio) at -20°C for ≥ 2 h in order to remove SDS, pelleted (14,000 rpm, 5 min), washed, air-dried, and resuspended in 6 M urea/2 M thiourea to solubilize proteins for enzymatic digestion. Protein concentration (typically 5-10 ug per case) was determined by the Bradford assay.

### Tryptic Digestion and LC-MS/MS Analysis

Protein extracts were reduced with 10 mM DTT (1 h, 37 °C), alkylated with 50 mM iodoacetamide (45 min, dark) and digested overnight at 37 °C with sequencing-grade trypsin (1:25 enzyme:protein) after diluting urea to < 1 M. Peptides were desalted using in-house C18 stage tips (3M Empore SPE-C18 disk, 47 mm, Sigma Aldrich, USA), dried and reconstituted in 2% acetonitrile/0.1% formic acid. Approximately 150 ng of peptides per sample were analyzed on a nanoElute (Bruker Daltonics) coupled to a timsTOF Pro mass spectrometer (Bruker Daltonics) with a CaptiveSpray source. Peptide separation was performed on a 15 cm × 75 µm ID C18 column (1.9 µm, Dr. Maisch) at 50 °C and 500 nL/min using a 5-35% acetonitrile linear gradient over 35 min. The instrument operated in data-dependent PASEF mode (1.1 s cycle). Data were processed using DataAnalysis 5.1 (Bruker Daltonics) to extract ion mobility-resolved spectra and mobility versus m/z distributions, as well as to determine the summed fragment ion intensities. Then, data were searched against the human proteome (UniProtKB) for protein identification and relative quantification.

### MS Data Analysis

MS spectra were analyzed using PEAKS Pro Studio v10.6 (Bioinformatics Solutions Inc.) against the human UniProtKB database. Search parameters included semi-specific trypsin, ≤2 missed cleavages, 15 ppm precursor mass tolerance, 0.02 Da fragment mass tolerance and a false discovery rate (FDR) <1% at peptide and protein levels. Proteins identified with ≥2 unique peptides and meeting FDR criteria were considered confidently identified. In selected cases, the PEAKS SPIDER algorithm was used to detect sequence variants and post-translational modifications not covered by the standard database search.

Each patient/sample was interpreted alongside an amyloid-negative control tissue of the same type. Data interpretation focused on enrichment of known amyloid-related proteins in the patient sample relative to control tissue. An amyloid-associated protein signature was considered present when at least two of the three universal amyloid-associated proteins, APOE, APOA4 and SAMP, showed peak areas ≥10⁵. Candidate fibril proteins were considered positive when they corresponded to a recognized amyloid fibril protein according to current International Society of Amyloidosis (ISA) nomenclature [13], exceeded the ≥10⁵ peak-area threshold and were enriched relative to the corresponding amyloid-negative control tissue. Cases showing an amyloid-associated protein signature without a confidently identified fibril protein were classified as indeterminate.

### Statistical Analysis

Diagnostic agreement between Congo red staining, immunohistochemistry (IHC), immunofluorescence (IF) and LC-MS/MS was assessed using contingency tables. For binary performance analyses, indeterminate (“doubtful”) classifications and unavailable results were excluded. Overall agreement was expressed as percentage concordance. Sensitivity and specificity were calculated with exact binomial 95% confidence intervals (95% CI) and inter-method agreement was assessed using Cohen’s kappa (κ) with 95% CI. LC-MS/MS was used as the reference method for these comparisons.

## Results

### Standardization and implementation of a workflow for amyloid subtyping in clinical practice

To evaluate and standardize a robust workflow for routine amyloid subtyping, we first analysed three positive control tissues: heart and tendon tissues with a known ATTR amyloid and a kidney sample with a confirmed AL amyloid (Figure 1A). In parallel, a panel of amyloid-negative control tissues (heart, kidney, adipose tissue, spleen, stomach, duodenum, liver, tongue, nerve, skin, prostate, testis, bladder, and gallbladder) was analysed to establish baseline expression levels of amyloidogenic and associated proteins. These controls served as reference samples to assess analytical specificity and to define quantitative criteria for fibril protein identification in cases with clinical suspicion of amyloidosis (Figure 2).

**Figure 1.**
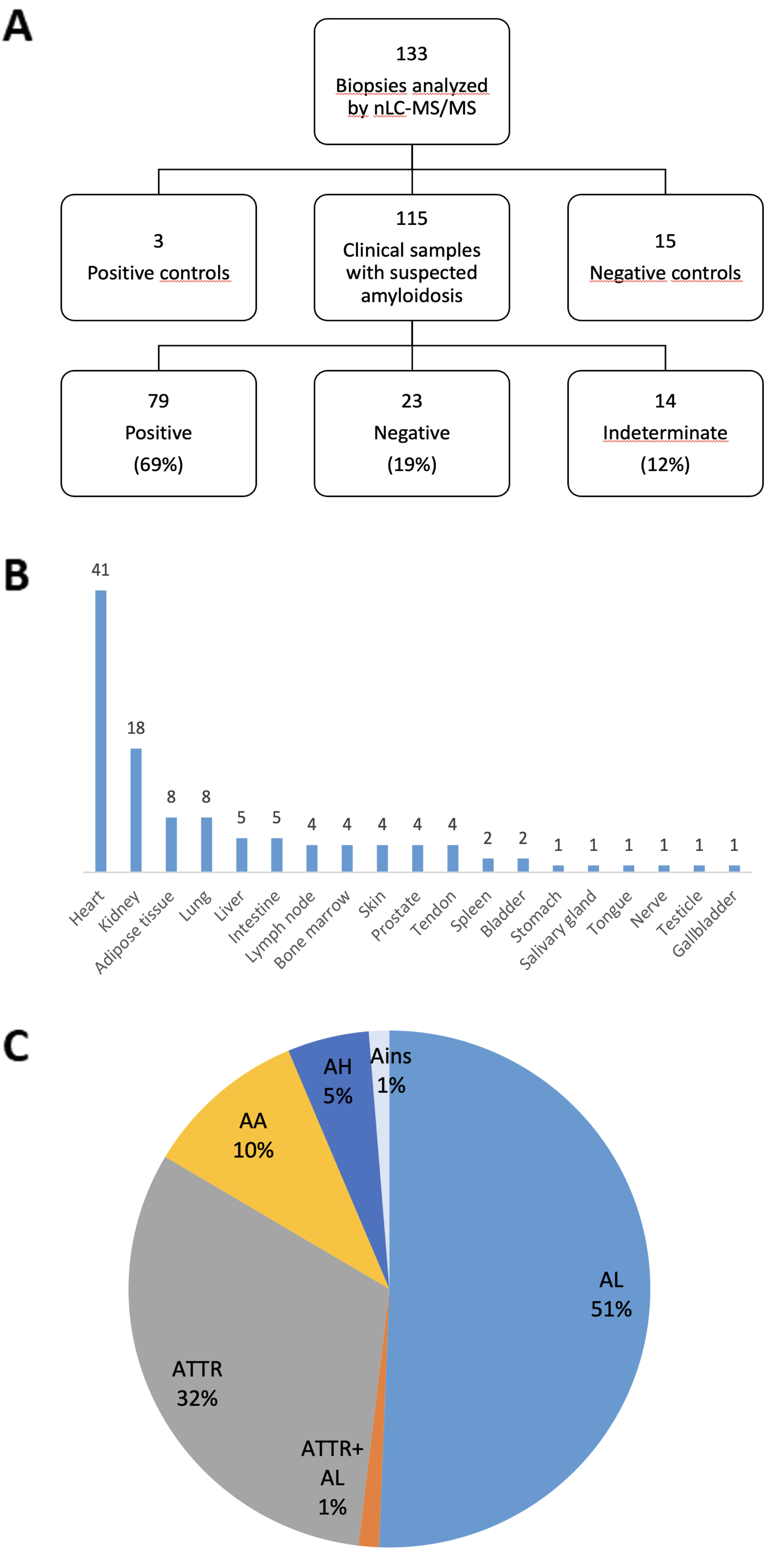
Overview of the prospective cohort and LC-MS/MS amyloid typing results. A) Flow diagram summarizing the 115 FFPE biopsies analyzed and their classification by LC-MS/MS. B) Distribution of biopsy tissue types included in the cohort. C) Distribution of amyloid subtypes among the 79 LC-MS/MS-positive cases.

**Figure 2.**
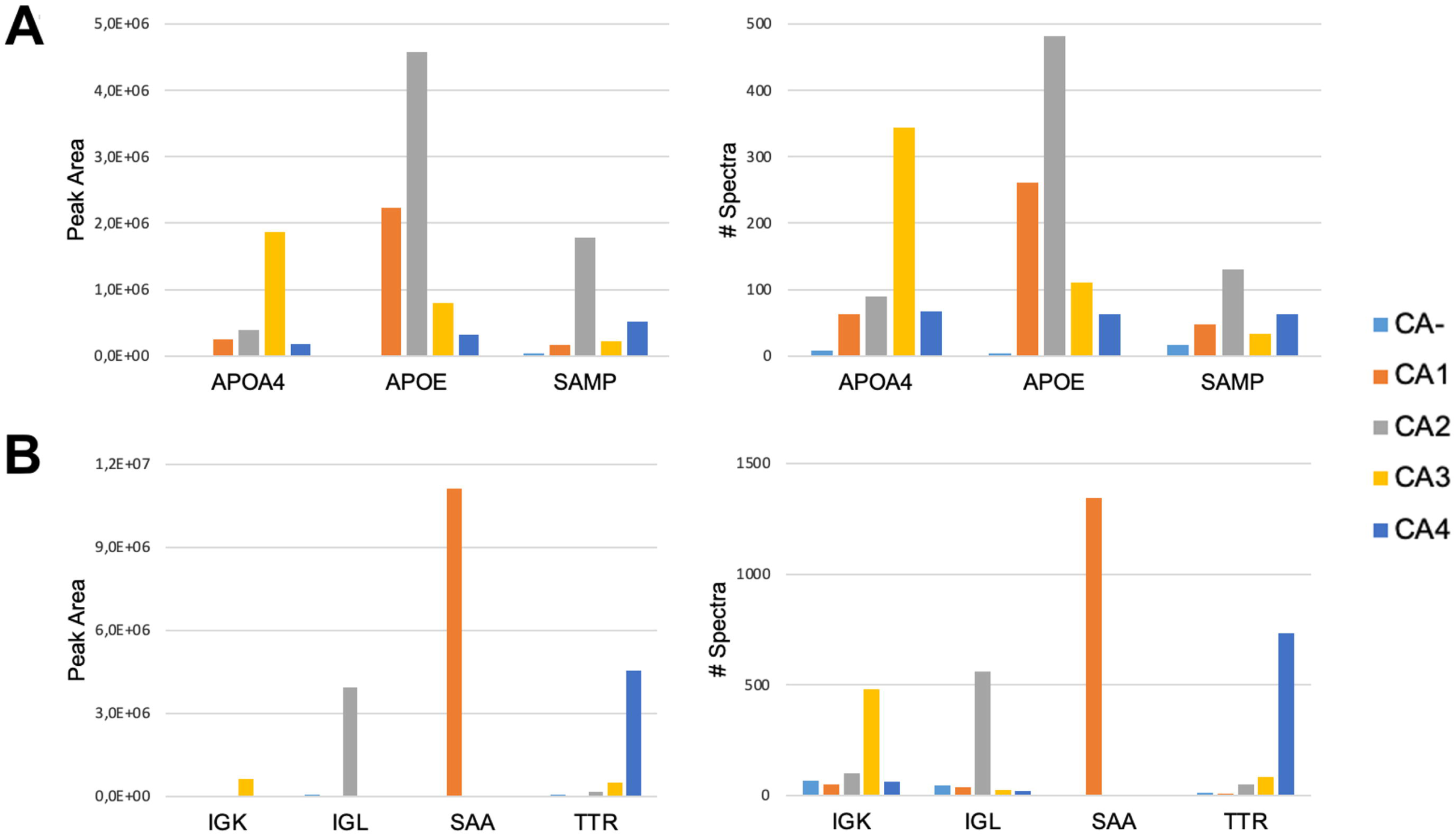
Standardization of the workflow for amyloid subtyping. Representative proteomic profiles from five cardiac biopsies, showing amyloidogenic proteins that were relatively quantified by peak area (left), while the number of spectra matching each protein served as a surrogate of abundance (right). A) Optimization of interpretation criteria. Threshold values (peak area ≥10⁵, PEAKS algorithm) were determined for nonspecific amyloid-associated proteins (APOE, APOA4, SAMP) using FFPE cardiac biopsy specimens. B) Amyloid subtyping. One patient without cardiac amyloidosis (CA-) was negative for all known amyloidogenic proteins, whereas four patients with suspected cardiac amyloidosis (CA1-CA4) showed distinct molecular signatures: AL κ-type, AL λ-type, AA, and ATTR. Each report was reviewed and validated by a pathologist in the context of clinical, immunohistochemical and imaging findings.

To empirically establish a quantitative threshold for the interpretation of amyloid-associated proteins, peak area values obtained with PEAKS software were examined in a calibration set comprising nine LC-MS/MS-positive and nine LC-MS/MS-negative FFPE cardiac biopsy specimens. A peak area of ≥10⁵ was selected as the working threshold for an amyloid-associated signal. APOA4 and APOE exceeded this threshold in all MS-positive specimens and remained below it in all negative specimens, whereas SAMP showed limited isolated overlap. Based on these observations, an amyloid-associated protein signature was considered present when at least two of the three universal amyloid-associated proteins (APOA4, APOE and SAMP) exceeded the 10⁵ peak-area threshold. Within this cardiac calibration set, the criterion correctly classified all 18 specimens (Fisher’s exact test, p = 4.1×10⁻⁵). For amyloid subtype assignment, the candidate fibril protein was additionally required to belong to the recognized list of amyloidogenic proteins, exceed the same quantitativethreshold and be enriched in the patient sample relative to the corresponding amyloid-negative control tissue (representative profiles are shown in Fig. 2A).

In all confirmed amyloid cases, hallmark amyloid-associated proteins such as APOA4, APOE and SAMP, were consistently detected, serving as internal indicators of amyloid deposition. If none of the known fibril proteins exceeded the quantitative threshold, and only these nonspecific amyloid-associated proteins were present, the result was classified as indeterminate or doubtful. Such findings typically reflected very small deposits or peptide signals below the confident identification level. Similarly, absence of the hallmark amyloid-associated proteins also resulted in indeterminate classification.

All MS results for each case were summarized in a concise report (Figure 2, Supplementary Table S1), listing the top proteins by spectral count, number of unique peptides and peak area, with the identified amyloidogenic protein highlighted. Reports were sent to the Pathology Department, where a pathologist reviewed and validated the proteomic interpretation in conjunction with clinical and histological data. After validation, a final diagnostic report was issued to the referring clinical department. The average response time from biopsy receipt to finalized report was approximately two weeks, compatible with clinical decision-making timelines.

### Overview of results obtained in the prospective study

From 2020 to 2025, 115 biopsy samples from patients with suspected amyloidosis were analyzed by LC-MS/MS (Figure 1A). The patients presented with diverse clinical syndromes, including cardiomyopathy, nephrotic syndrome and peripheral neuropathy, in which amyloidosis was considered in the differential diagnosis. Samples originated multiple tissues (Figure 1B), with a predominance of cardiac biopsies (36%), followed by kidney (16%), adipose tissue (7%), lung (7%), liver (4%), intestine (4%). Less frequent sites included lymph node, bone marrow, skin, prostate and tendon (each 3.5%), with other tissues represented by ≤2 cases. Of the 115 cases analyzed, 79 (69%) were confirmed positive for amyloid by MS, meaning that a specific fibril protein was identified. In 22 cases (19%), no amyloidogenic proteins were detected, and the samples were classified as amyloid-negative (Figure 1A). These negative results led to a review of the histology: the discordant Congo red-positive/LC-MS/MS-negative cases included histologically ambiguous or highly focal deposits and, in some instances, possible false-positive Congo red staining. The remaining 14 cases (12%) produced indeterminate results. These typically showed the presence of general amyloid-associated proteins (APOE, APOA4, SAMP) without a fibril protein exceeding the positivity threshold, likely reflecting very limited amyloid content. Such cases were reported as “amyloid present, subtype inconclusive” and generally corresponded to minimal or histologically ambiguous deposits. In some instances, the opposite pattern was observed: detection of amyloidogenic proteins, particularly Ig light chains (e.g., IGK), in the absence of a complete amyloid signature.

The proteomic analysis revealed a diverse spectrum of amyloid subtypes within the cohort (**Figure 1C**). AL amyloidosis was the most frequent subtype, identified in 40 cases (51%), including 23 lambda (AL λ-type) and 17 kappa (AL κ-type) light-chain cases, consistent with the known predominance of λ chains in AL amyloidosis. ATTR amyloidosis (transthyretin) was the second most common subtype, detected in 25 cases (32%). These included both wild-type (age-related systemic) and hereditary forms associated with pathogenic TTR variants. AA amyloidosis, caused by serum amyloid A protein, was confirmed in 8 cases (10%), typically in patients with chronic inflammatory diseases. Together, AL, ATTR, and AA amyloidosis represented the vast majority of diagnoses in the cohort.

### Identification of rare and mixed amyloid types

Beyond the common subtypes (AL, ATTR and AA), four cases of immunoglobulin heavy-chain (AH) amyloidosis were identified (Figure 1C). We also detected a case of localized, iatrogenic amyloidosis: an AIns amyloid deposit at insulin injection sites in a diabetic patient. The patient presented with subcutaneous nodules that, upon MS analysis, contained insulin as the dominant protein (peak area > 10^7^; 1433 spectra), accompanied by the universal amyloid-associated proteins APOE, APOA4, and SAMP (each > 10^5^) (Figure 3A). This profile confirmed insulin as the causative protein in the localized amyloid.

**Figure 3.**
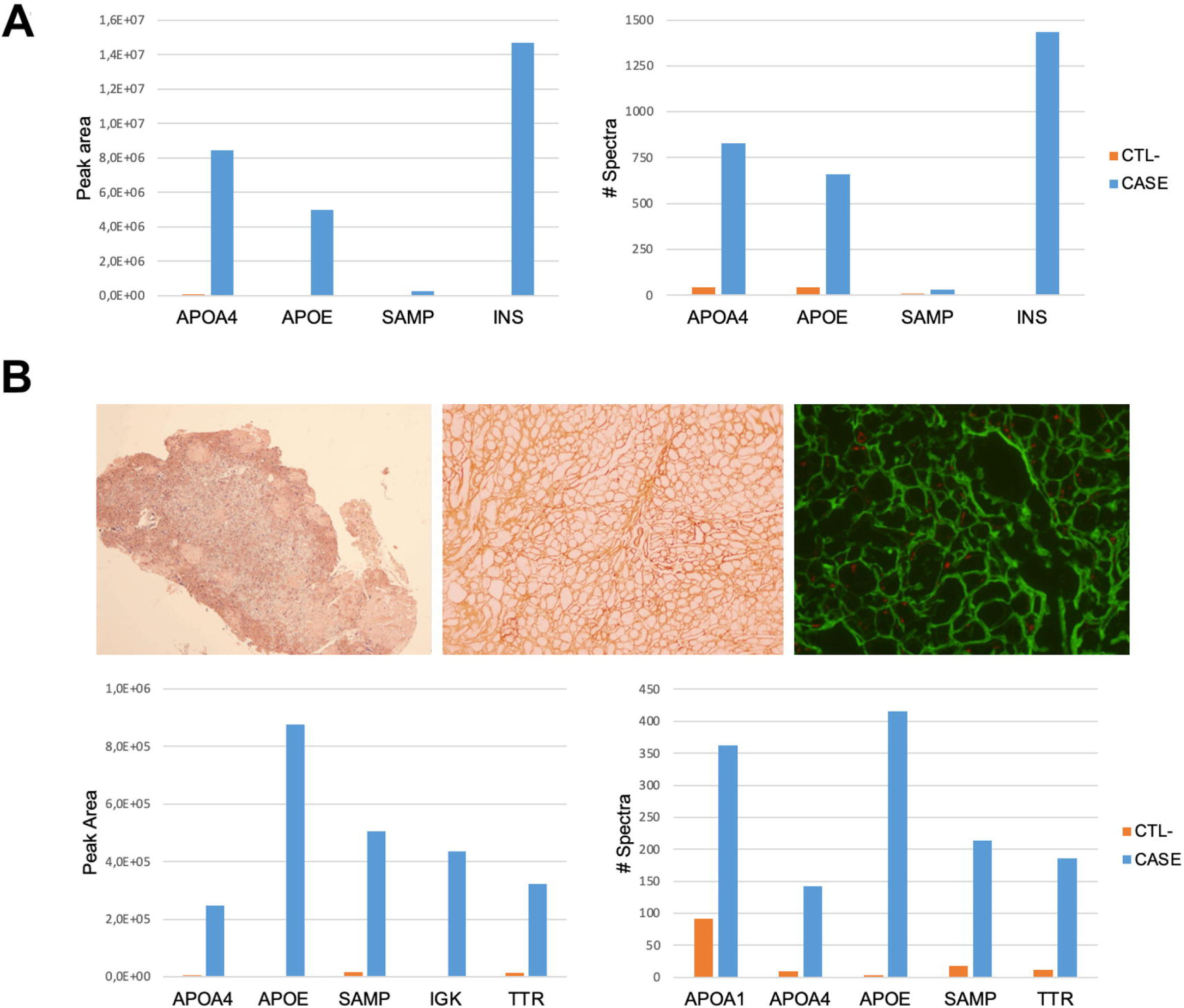
Identification of rare and mixed amyloid subtypes. A) MS identification of iatrogenic amyloidosis. Peak area (left) and peptide spectral count profiles (right) for an injection-site amyloid deposit. INS, insulin. B) Cardiac biopsy with mixed amyloidosis. Upper panel: Congo red birefringence under cross-polarized light, immunohistochemistry for transthyretin, and immunofluorescence for κ light chains. Bottom panel: peak area and peptide spectral count profiles showing concurrent detection of IGK and TTR by LC-MS/MS, consistent with mixed amyloidosis.

A mixed ATTR+ALκ amyloid deposit was identified in one cardiac biopsy (Figure 3B). Histological evaluation demonstrated Congo red-positive deposits, while IHC showed transthyretin reactivity and IF detected κ light chains. LC-MS/MS demonstrated concurrent enrichment of TTR and IGK, both exceeding the quantitative threshold, supporting the presence of mixed ATTR and ALκ amyloidosis. The patient had initially been managed under a diagnosis of ATTR amyloidosis and was on treatment with tafamidis: identification of a coexisting additional ALκ component prompted diagnostic reassessment, including hematologic evaluation and follow-up.

### Proteomic characterization of amyloid deposits beyond subtype identification

Comprehensive LC-MS/MS analysis also enabled the molecular characterization of amyloidogenic proteins, including the detection of pathogenic mutations and post-translational modifications (PTMs) directly from tissue deposits.

In one cardiac ATTR case carrying TTR c.148G>A (p.Val50Met; historically p.Val30Met), MS detected the variant peptide and, using PEAKS SPIDER, estimated the relative abundance of variant and wild-type peptides (Figure 4A). In one AA case, exploratory SPIDER analysis identified a markedly more complex SAA modification profile than in an amyloid-negative control, including oxidation, deamidation and mass shifts consistent with glycation (Figure 4B). These observations illustrate additional molecular information obtainable from tissue proteomics but were not intended as validated diagnostic endpoints.

**Figure 4.**
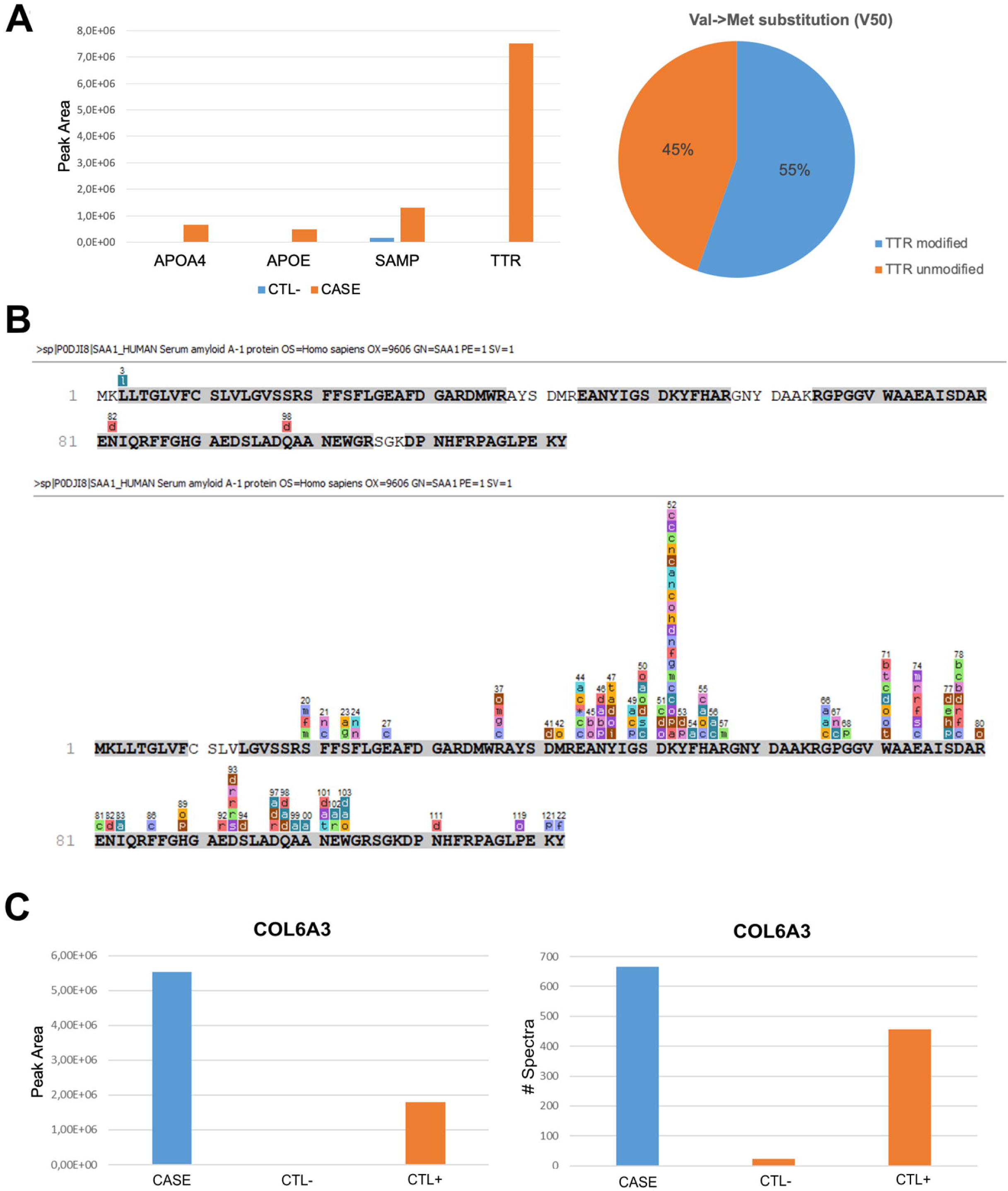
Molecular characterization of amyloidogenic proteins by LC-MS/MS. A) Mass spectrometric identification of ATTRv amyloidosis (p.Val50Met) in a cardiac biopsy. Transthyretin (TTR) dominated the protein content (peak area > 10⁶) (left panel) and the SPIDER algorithm enabled the detection and quantification of variant and wild-type peptides (right panel). B) Post-translational modification profiling of serum amyloid A (SAA) in a cardiac biopsy. The top panel shows the sequence coverage map of SAA in a negative control sample (AA-), while the AA+ sample (bottom panel) exhibited >100 modified peptides, including oxidation, deamidation, and glycation events (colored annotations indicate modified residues). C) Enrichment of the extracellular matrix protein COL6A3 (in terms of Peak Area and Spectral counting), in one renal amyloid deposit (blue bar) compared with two matched control tissues (orange bars).

Extracellular matrix proteins were also enriched in selected deposits; COL6A3 was prominent in a subset of renal amyloid biopsies compared with matched controls (Figure 4C), suggesting tissue-specific remodeling associated with amyloid deposition.

### Cross-Method Validation and Correlation

Agreement heatmaps including positive, negative and doubtful categories are shown in Figure 5A. Discordances occurred mainly in Congo red-positive/MS-negative specimens and doubtful histological cases, many of which were classified as negative by MS.

**Figure 5.**
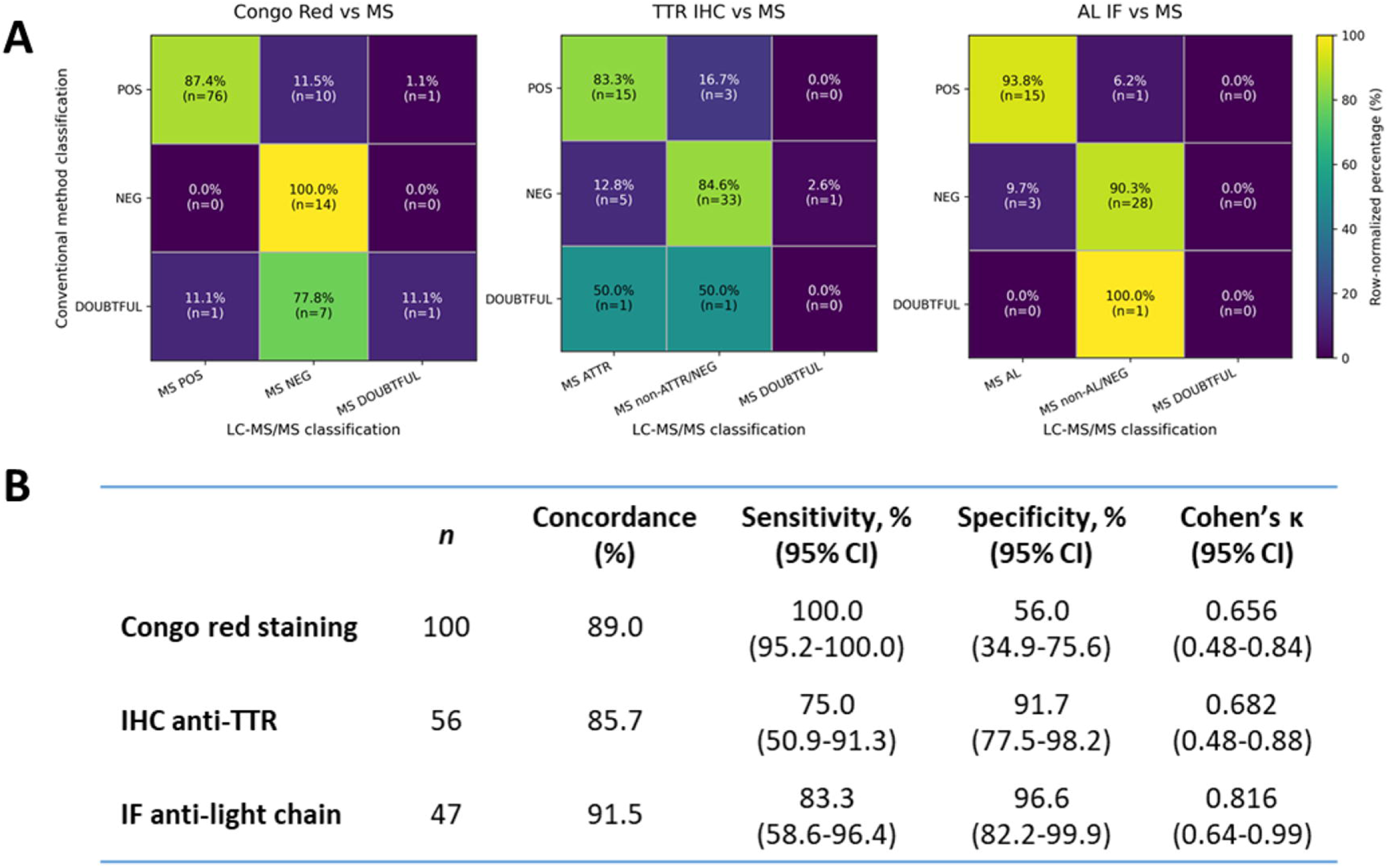
Agreement and diagnostic performance of conventional methods compared with LC-MS/MS. A) Agreement matrices comparing Congo red staining, anti-TTR immunohistochemistry, and AL light-chain immunofluorescence with LC-MS/MS. Heatmaps display row-normalized percentages with absolute case numbers indicated in each cell; positive, negative, and indeterminate (“doubtful”) categories are shown. B) Summary of binary diagnostic performance after exclusion of indeterminate/doubtful and unavailable results, including concordance, sensitivity, specificity, and Cohen’s κ with 95% confidence intervals. LC-MS/MS was used as the reference method. For Congo red staining, MS positivity referred to detection of any amyloidogenic fibril protein; for anti-TTR IHC and anti-AL IF, MS positivity referred to the corresponding ATTR and AL subtypes, respectively.

When all three categories were considered, concordance was 82.7% (91/110) for Congo red, 81.4% (48/59) for anti-TTR IHC, and 89.6% (43/48) for AL IF. In binary analyses excluding doubtful cases (Figure 5B), concordance was 89.0% for Congo red (sensitivity 100.0%, specificity 56.0%, κ=0.656; 95% CI 0.48–0.84), 85.7% for anti-TTR IHC (75.0%, 91.7%, κ=0.682; 0.48–0.88), and 91.5% for AL IF (83.3%, 96.6%, κ=0.816; 0.64–0.99). LC-MS/MS provided additional diagnostic information in 64/110 evaluable cases (58.2%): 53/110 (48.2%) represented additional subtyping information (42 without available IHC/IF-based subtype assignment, three indeterminate antibody-based results, and eight alternative, rare, or mixed profiles), while 11 were Congo red-positive/MS-negative diagnostic discordances.

## Discussion

We prospectively implemented an LC-MS/MS workflow for amyloid typing at Hospital Universitario A Coruña (HUAC), analyzing 115 consecutive FFPE biopsies from patients with suspected systemic or localized amyloidosis between 2020 and 2025. Despite the absence of routine laser microdissection. LC-MS/MS identified a definitive fibril protein in 79/115 cases (69%), with AL and ATTR as the predominant subtypes and additional AA, AH, mixed ATTR+AL and rare hereditary or localized forms also identified, supporting the feasibility of this approach in routine clinical practice.

Overall agreement between conventional techniques and LC-MS/MS was high, although proteomics provided clinically relevant information beyond that obtainable from routine histological and antibody-based methods [8]. Among conventional techniques, IF showed the highest agreement with MS classification, whereas Congo red staining, despite its high sensitivity, showed lower specificity because of discordant Congo red-positive/LC-MS/MS-negative cases.

Importantly, LC-MS/MS provided additional diagnostic information in 64/110 evaluable cases (58.2%). In 53 cases (48.2%), this represented additional amyloid subtyping information: 42 lacked an available IHC/IF-based subtype assignment, 3 remained indeterminate after antibody-based testing and eight corresponded to alternative fibril proteins or mixed deposits not resolved by the available conventional methods. A further 11 cases showed discordant Congo red-positive/LC-MS/MS-negative results, possibly reflecting nonspecific Congo red staining, highly focal deposits, differences between serial tissue sections, loss of amyloid-containing material during processing or insufficient amyloid for proteomic detection. Together, these findings support the complementary value of LC-MS/MS, particularly for FFPE specimens, indeterminate antibody results and unexpected amyloid types.

A major advantage of MS is its sequence-based identification of fibril proteins, broadening diagnostic scope beyond that achievable with limited antibody panels and enabling characterization of the molecular composition of amyloid deposits [14,15]. In our cohort, LC-MS/MS identified rare forms of amyloidosis that may be difficult to establish using routine IHC when validated antibodies are unavailable: identification of hereditary ATTRv led to genetic evaluation, while identification of localized AIns amyloidosis supported modification of insulin injection practices. MS was also particularly valuable in complex samples containing mixed amyloid deposits. The coexistence of AL and ATTR amyloid within the same specimen has been previously described [16]. In our cohort, LC-MS/MS identified a cardiac biopsy with co-existing ATTR and ALκ amyloid, illustrating the ability of proteomic analysis to resolve dual amyloid pathology that antibody-based methods alone may fail to interpret. This distinction is clinically relevant given that the two components may require different therapeutic strategies [16].

Consistent with previous reports [17–20], our findings suggest that Congo red-guided FFPE sections containing sufficient amyloid can provide interpretable results without routine laser microdissection. The use of a defined peak-area cutoff (≥10⁵), amyloid-associated proteins and matched amyloid-negative tissue controls provided a consistent framework for distinguishing candidate fibril proteins from background signals, conceptually aligned with recent quantitative strategies assessing amyloid precursor enrichment relative to non-amyloid tissue [20,21]. Nevertheless, our threshold was empirically derived from a small internal calibration set restricted to cardiac biopsies and subsequently applied across different tissue types. Its analytical performance therefore warrants confirmation in larger independent cohorts and, where feasible, tissue-specific validation sets.

Importantly, the workflow was implemented without routine laser microdissection (LMD). Although LMD remains widely used for proteomics-based amyloid typing of FFPE tissue, alternative strategies including manual microdissection, macrodissection, whole-tissue analysis and tissue-homogenization workflows have also been successfully applied [19,20,22]. Recent expert reviews emphasize that both approaches can support amyloid typing when appropriately validated [23]. Our findings extend this evidence by demonstrating the feasibility of a non-LMD workflow across routine FFPE specimens from multiple tissue types in a prospective clinical setting. Nevertheless, microdissection may remain advantageous in specimens with sparse or spatially heterogeneous deposits, substantial blood contamination or suspected dual amyloidosis. Several practical limitations should be considered. Reliable MS typing more generally depends on adequate amyloid content and very small, heterogeneous or degraded specimens may yield indeterminate results. Appropriate instrumentation, experienced personnel, bioinformatic infrastructure and robust quality-control procedures are also required. Centers without in-house capabilities may therefore benefit from collaboration with reference laboratories or regional MS hubs. Although MS is more expensive than conventional staining, accurate subtype identification may prevent misclassification and inappropriate treatment in a disease in which therapeutic decisions depend directly on correct typing.

Beyond subtype identification, LC-MS/MS provided additional molecular information. In ATTR amyloidosis, MS enabled discrimination between variant (ATTRv) and wild-type (ATTRwt) forms according to current ISA nomenclature [13]. Because more than 100 amyloidogenic TTR variants have been described [24], detection of a variant peptide by MS should be confirmed by genetic sequencing, and peptide-level quantification of variant versus wild-type TTR, while investigational, could provide additional biological information pending validation as a predictive or pharmacodynamic biomarker. In AA amyloidosis, exploratory tandem-MS analysis revealed a more complex pattern of oxidation, deamidation and glycation-related modifications across the SAA sequence than in control tissue. Such modifications could alter protein conformation or epitope accessibility and contribute to weak or discordant IHC findings [25], potentially through advanced glycation end products and RAGE-related pathways [26]. Given the exploratory nature of this analysis, these observations should be regarded as hypothesis-generating. We also observed enrichment of extracellular matrix and basement membrane proteins, particularly COL6A3 in renal biopsies, consistent with proteomic studies suggesting that matrix-remodeling proteins reflect tissue-specific responses to amyloid deposition rather than components of the fibril core itself [15]. Implementation of MS-based amyloid typing required close collaboration between pathology and proteomics, consistent with the view that translating MS-based proteomics into molecular pathology requires analytical validation, standardized quality-control procedures and integration within established diagnostic workflows [27]. At HUAC, pathologists selected Congo red-positive tissue for analysis and proteomic results were interpreted in clinicopathological context before reporting, with turnaround times of 2 weeks that are compatible with clinical decision-making, and urgent analyses achievable within 2-3 days. Our findings are consistent with previous large-scale clinical experiences that have established MS-based proteomics as a reference method for amyloid typing, including the Mayo Clinic series of more than 16,000 clinical samples [1], the UK National Amyloidosis Centre experience [9] and a French nationwide cohort spanning 10 years [28]. However, these reports largely describe accumulated or retrospective diagnostic experience. To our knowledge, the present study is the first prospective series evaluating the integration of LC-MS/MS-based amyloid typing of routine FFPE biopsies into the diagnostic workflow of a tertiary hospital, and the first such experience reported from Spain, applied over five years across multiple tissue types through direct coordination between pathology and proteomics services. The translational contribution of this approach is summarized in Table 1.

**Table 1.**
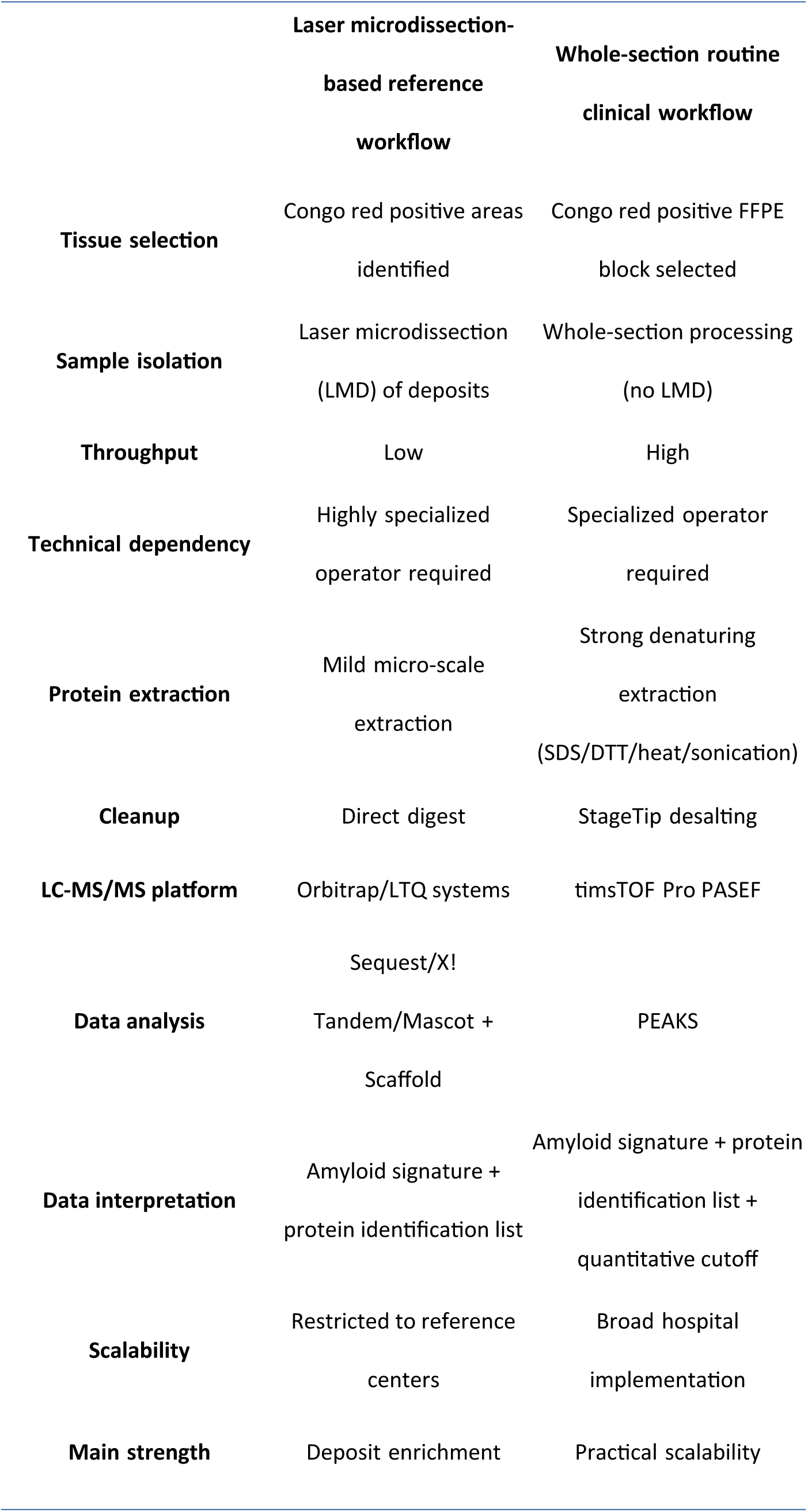
Comparison of a typical LMD-based amyloid typing workflow with the non-LMD routine clinical workflow implemented at HUAC. The comparison summarizes general characteristics of commonly used LMD-based approaches and the workflow evaluated in the present study.

In conclusion, LC-MS/MS amyloid typing was feasible within our routine FFPE workflow and complemented conventional pathology by resolving common, rare and mixed amyloid types. Close integration of pathology and proteomics enabled the generation of clinically useful reports, while sequence-level analysis provided additional molecular information. These findings support the implementation and further validation of proteomic amyloid typing in hospital-based diagnostic practice.

## Data Availability

All data produced in the present study are available upon reasonable request to the authors

## Acknowledgments

The authors would like to thank the staff of the Department of Pathology at Hospital Universitario A Coruña for their technical support in tissue selection and sample processing. The authors also thank Nieves Domenech for her valuable role in promoting the collaboration between the Proteomics Unit and the Department of Cardiology.

Part of the protein analysis was performed by the ICTS “NANBIOSIS”, specifically by the Proteomics Unit of CIBER-BBN at SERGAS (Spain).

## Data availability statement

Proteomics data are available from the corresponding author upon reasonable and justified request, subject to institutional requirements and compliance with applicable data protection regulations.

## Declaration of generative AI use

During the preparation of this manuscript, the authors used ChatGPT (OpenAI) for English-language editing, condensation, and readability, and for the generation of the graphical abstract. All AI-assisted text and graphical content were reviewed and verified against the underlying data and results by the authors, who take full responsibility for the content of the manuscript.

---

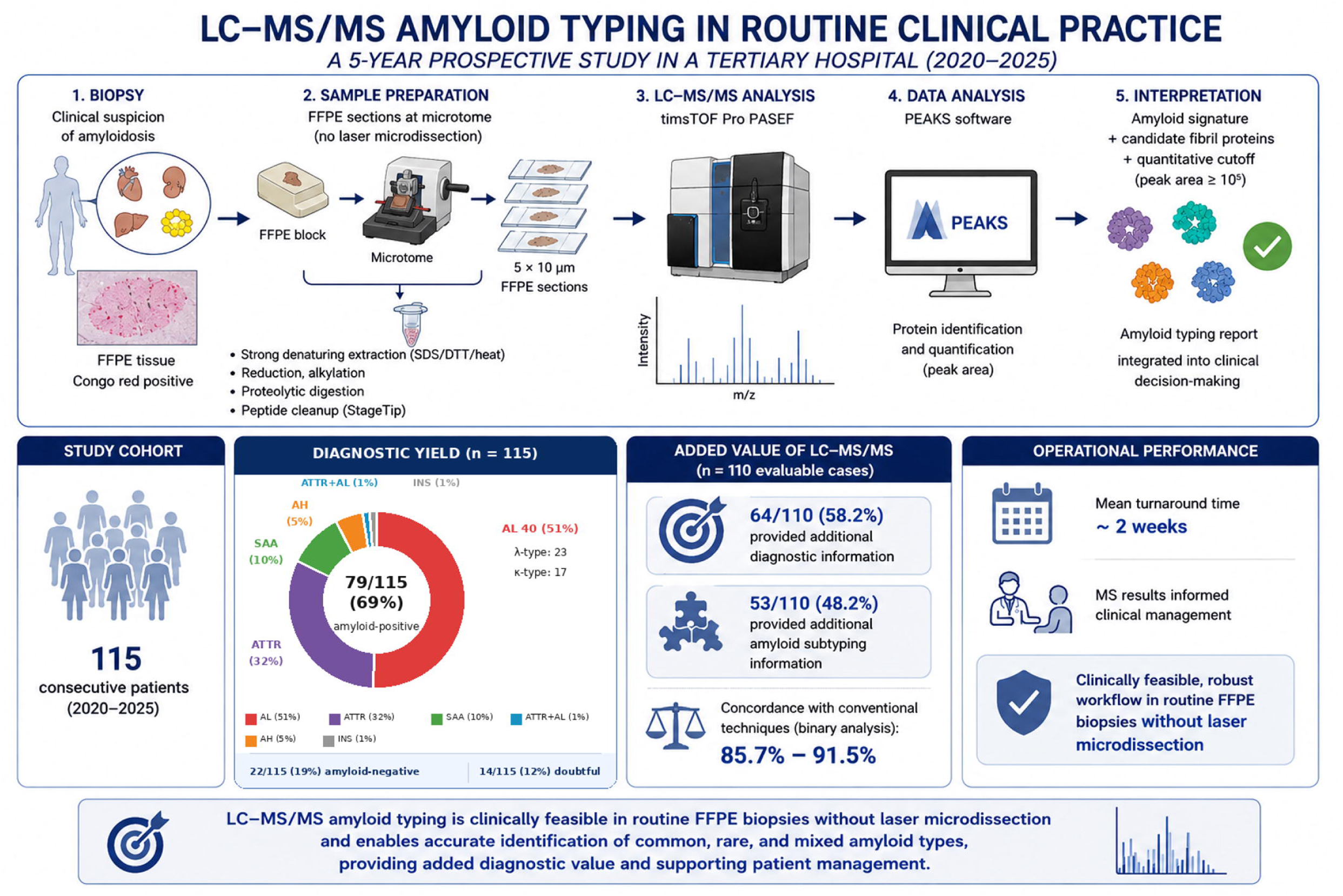

